# Molecular profiling of repeated self-sampled blood reveals dynamic immune phenotypes in young adults

**DOI:** 10.64898/2026.01.16.26344184

**Authors:** Annika Bendes, Sophia Björkander, Maura M. Kere, Simon Kebede Merid, Ashish Kumar, Leo Dahl, Zhebin Yu, Amelie Vogt, Changil Kim, Qiang Pan-Hammarström, Anna Bergström, Inger Kull, Anne-Sophie Merritt, Sandra Ekström, Alexandra Lövquist, Ben Murrell, Niclas Roxhed, Erik Melén, Jochen M. Schwenk

## Abstract

Population-based studies of circulating blood proteins have provided profound insights into human biology. However, short-lived changes often remained undetected. To address this, we performed a comprehensive longitudinal dried blood spot (DBS) self-sampling study in 808 young adults of the BAMSE cohort during 2020-2022. We profiled serological, autoimmune, and proteomic phenotypes in relation to SARS-CoV-2 exposures (infection, vaccination), physiological traits, genetic variation, and blood counts. Data-driven seroclustering revealed dynamic immune response to both exposures, while analysis of anti-interferon autoantibodies (AAbs) uncovered associations of pre-existing stable AAbs with prolonged COVID-19 symptoms. Genome-wide mapping determined 664 pQTLs, with cis-pQTLs associated showing increased longitudinal stability. We also identified relationships between blood cell counts and DBS proteins beyond the hematocrit effects. Paired pre- and post-exposure highlighted transient alterations for infection (e.g. LAP3) and vaccinations (e.g. TIMP3). Multi-molecular phenotyping in self-sampling can capture dynamic immune trajectories, informing precision medicine efforts with clinically valuable insights on short-term variability in health phenotypes.

## Introduction

Recent advances in studying molecular phenotypes have provided new insights into human health at population level ^1–3^. However, clinical outcomes can often be altered by short terms exposures, disproportionately affecting those with pre-existing health conditions. Hence, baseline characteristics and continuous insights are needed to interpret pre-existing, temporal, and acquired molecular phenotypes.

Understanding the landscape of molecular phenotypes in the general and younger population will add to informing future precision medicine approaches on how to treat patients of the future. As young adults become older, they are set to require care later in life. Thus, understanding and managing the relation of existing, shorter, transient or acquired health conditions will support countermeasures against upcoming chronic diseases. To enable large-scale monitoring in a more patient-centric manner, repeated self-sampling of blood has become increasingly attractive. The use of alternative liquid biopsies, such as dried blood spots (DBS), facilitates democratized access to molecular data without requiring in-clinic visits.

Establishing these novel sampling methods for multi-parameter molecular profiling remains the subject of current efforts to ensure accuracy and reproducibility. Among the accessible molecular phenotypes, circulating blood proteins offer attractive insights into genetics, environmental exposures, comorbidities, and treatment effects, reporting on a broad range of biological processes^4^. Immunoglobulins, for example, are well-established markers of immunological memory and acquired anti-SARS-CoV-2 antibodies (Abs) persist for up to a year^5^, and infections have also been linked to the onset of autoantibodies (AAbs), while other changes in the circulating blood proteome^6^, like cytokines^7^ and immune cell receptor activation^8^ are more short-lived. Compared to pathogen-specific Abs, inflammatory proteins may have been activated by different exposures and at different timepoints in a person’s life. Integrating both immune and proteomic profiling is therefore informative to capture the host response to infection, providing insights into immune variability, risk stratification, and persisting health consequences of infections and vaccinations.

To democratize health monitoring, self-sampling of dried blood spots (DBS) has become increasingly valuable. The use of DBS as an alternative liquid biopsy simplified access to molecular data without in-clinic visits. Benchmarking these novel methods for multi-parameter immune profiling against established routines is the subject of current efforts to ensure their accuracy and reproducibility^9–11^.

Here, we performed a comprehensive longitudinal DBS self-sampling study in 800 young adults of a population-based birth cohort to provide insights into immune resilience and susceptibility in young adults. By combining multi-analyte serology, autoimmune and proteomic data with genetic, clinical, and health history data, we reveal transient and persistent trajectories, highlighting molecular phenotypes before, during, and after SARS-CoV-2 exposures. Collectively, our results establish an actionable framework for remote molecular health monitoring for mechanistic insights and adjustable therapeutic management.

## Results

We studied self-sampled blood from young adults to investigate the impact of SARS-CoV-2 infections and vaccinations on their molecular phenotypes (Fig. 1a). The goal was to evaluate the feasibility of deconvoluting how molecular health information is impacted by pandemic exposures and how these shape dynamic multi-analyte phenotypes of the participants.

**Fig. 1:**
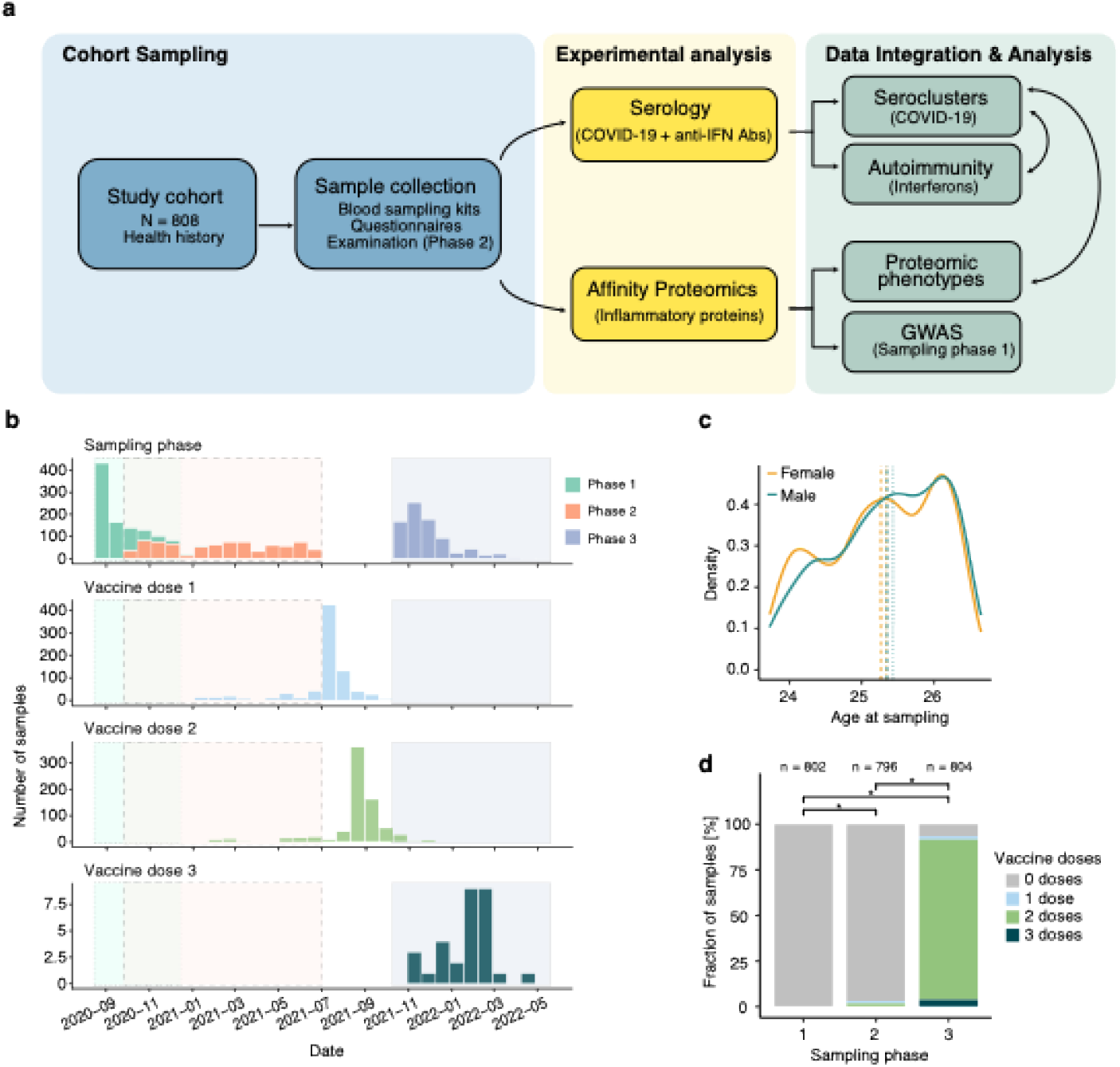
Study design for trans-pandemic analysis of dried blood spots. (a) Study design where 808 participants were asked to give dried blood spots at three different time points (sampling phases) and answer questionnaires. At sampling Phase 2, the participants had a clinical examination. The dried blood spot samples were then used for serological analyses measuring antibodies against COVID-proteins and human interferons, and proteomic analyses measuring 365 inflammatory proteins. (b) Samples were taken between August 2020 and February 2022, and Phase 3 was predominantly sampled after two vaccine doses. (c) Age distribution of males and females per sampling phase (vertical dashed lines) (d) Distribution of vaccinations per sampling phase. Asterisks represent FDR-adjusted p-values below 0.05 from Fisher’s exact test.

### Study design and cohort population

From the BAMSE birth cohort, 808 participants (64% females) completed a longitudinal study consisting of three blood self-sampling phases from August 2020 to May 2022 (Fig. 1b)^12^. In addition, participants answered health-related questionnaires for Phase 1-3, and in Phase 2, participants also underwent a clinical examination. There were no differences in age between the sexes, but a larger proportion of females smoked cigarettes, and males had higher BMI while females had higher body fat percentage (Fig. 1c, Tab. 1). Of the 808 participants, 791 (97.9%) successfully provided one quantitative DBS sample per phase. In Phase 1, no participants were vaccinated, 3.1% had received at least one dose by Phase 2, and 93.4% by Phase 3 (Fig. 1d). In Phase 1, 1.4% reported PCR-confirmed infections, 9.7% in Phase 2, and 18.9% in Phase 3 (Fig. S1, Tab. 1), where the Omicron wave added most new cases.

**Table 1.** Overall demographics of the 808 subjects in the study population^1,2^.

|  | Phase 1 |  |  |  | Phase 2 |  |  |  | Phase 3 |  |  |  |
| --- | --- | --- | --- | --- | --- | --- | --- | --- | --- | --- | --- | --- |
|  | All | Females | Males | *p (sex)* | All | Females | Males | *p (sex)* | All | Females | Males | *p (sex)* |
| **n** | 808 | 517 (64.0) | 291 (36.0) |  | 808 | 517 (64.0) | 291 (36.0) |  | 808 | 517 (64.0) | 291 (36.0) |  |
| **Age (years)** | 25.4<br>(23.7-26.7) | 25.3<br>(23.7-26.5) | 25.4<br>(23.7-26.7) | 0.141 | 25.9<br>(23.9-27.3) | 25.8<br>(23.9-27.3) | 25.9<br>(24.0-27.2) | 0.179 | 26.6<br>(24.9-27.8) | 26.5<br>(24.9-27.8) | 26.6<br>(24.9-27.7) | 0.165 |
| **Smoking (yes)** | 112 (13.9) | 83 (16.1) | 29 (10.0) | 0.019 | 88 (10.9) | 68 (13.2) | 20 (6.9) | 0.007 | 81 (10.1) | 60 (11.7) | 21 (7.3) | 0.051 |
| **BMI category** | - | - | - | - |  |  |  | <0.001 | - | - | - | - |
| **<25.0** |  |  |  |  | 603 (75.4) | 406 (79.8) | 197 (67.7) |  |  |  |  |  |
| **≥25.0** |  |  |  |  | 197 (24.6) | 103 (20.2) | 94 (32.3) |  |  |  |  |  |
| **BMI (kg/m<sup>2</sup>)** | - | - | - | - | 22.6<br>(14.8-59.9) | 22.1<br>(14.8-59.9) | 23.5<br>(16.8-48.3) | <0.001 | - | - | - | - |
| **Body fat percentage** | - | - | - | - | 23.4<br>(5.9-51.1) | 26.1<br>(11.6-51.1) | 17.5<br>(5.9-43.8) | <0.001 | - | - | - | - |
| **SmiNet case (yes)<sup>3</sup>** | 11 (1.4) | 7 (1.4) | 4 (1.4) | 1 | 78 (9.7) | 53 (10.3) | 25 (8.6) | 0.535 | 152 (18.9) | 104 (20.2) | 48 (16.5) | 0.223 |
| **Vaccinated COVID-19 (yes)<sup>4</sup>** | 0 (0.0) | 0 (0.0) | 0 (0.0) |  | 25 (3.1) | 17 (3.3) | 8 (2.8) | 0.833 | 753 (93.4) | 482 (93.6) | 271 (93.1) | 0.883 |
| **Number of vaccine doses** | - | - | - | - |  |  |  | 0.613 |  |  |  | 0.817 |
| **1 dose only** |  |  |  |  | 12 (1.5) | 7 (1.4) | 5 (1.7) |  | 17 (2.1) | 12 (2.3) | 5 (1.7) |  |
| **2 doses only** |  |  |  |  | 13 (1.6) | 10 (1.9) | 3 (1.0) |  | 708 (87.8) | 454 (88.2) | 254 (87.3) |  |
| **3 doses** |  |  |  |  |  |  |  |  | 28 (3.5) | 16 (3.1) | 12 (4.1) |  |

Continuous variables were analysed by the Mann-Whitney U-test and are displayed as median with range. Dichotomous variables were analysed by the Fisher's exact test and are displayed as number and percentage. <sup>1</sup>All subjects with at least one approved serology- or Olink-measurement from at least one phase. <sup>2</sup>One female subject lacks smoking status in Phase 1, one male subject lacks smoking status in Phase 2, eight female subjects lack BMI and body fat percentage at Phase 2, two females and three males lack smoking status in Phase 3, two female subjects lack SmiNet case- and COVID-19 vaccination information in Phase 3 due to lacking date of filter paper sampling. <sup>3</sup>SmiNet (a national register for notifiable transmittable diseases in Sweden, covering all positive PCR tests (Public Health Agency of Sweden. SmiNet. In Swedish. Available at: https://www.folkhalsomyndigheten.se/sminet)). A SmiNet-case was defined as a positive entry date in SmiNet preceding the date of DBS filter paper sampling in each respective phase. <sup>4</sup>Information on vaccination against COVID-19 was obtained by linkage to the National Vaccination Register. COVID-19 vaccination was defined as receiving the vaccination prior to the date of DBS filter paper sampling in each respective phase.

### Classification of antibody response by multi-analyte serology

To assess the antibody response against SARS-CoV-2 over time, we performed multi-analyte serology analysis of circulating human IgG antibodies in DBS samples using our previously validated protocol^13^. Based on population-based cutoffs, 13.9% were N-seropositive (N+) in sampling Phase 1, 22.2% in Phase 2, and 26.0% in Phase 3 (averaged over several N proteins). The average S-seropositivity (S+) increased from 15.1% to 28.8%, to 97.3% across the sampling phases, reflecting infection and vaccination status. Similarly, RBD seropositivity (RBD+) increased from 9.5%, 19.8% to 96.8%. This data follows figures by the public health authorities on infection and vaccination rates^14^ and previous studies in a healthcare setting^15^.

### Seroclustering of antibody response by multi-analyte serology

To capture the heterogeneity of the immune response, considering intermediate or partial responders, we used data-driven clustering to stratify the samples into four distinct seroclusters based on serology (Fig. 2A), confirming previous insights^16^. As shown in Fig. 2a-d, Serocluster 1 consisted predominantly of seronegative samples (N-, S-, and RBD–), while Serocluster 2 had an average of 96.9% S+, but slightly lower positivity rates for N+ (89.4%) and RBD+ (74.2%). RBD+ rates were lower for the omicron variants BA.1 and BA2 (21.2%), compared to the earlier RBD versions (89.4%). Serocluster 3 contained ≥90% of seropositive samples with an average of 84.6% N+, of which all were deemed S+ and RBD+. All samples in Serocluster 3 were seropositive for both omicron variants. Lastly, Serocluster 4 was primarily S+ (100%) and RBD+ (99.9%), with only 3.2% being N+.

**Fig. 2:**
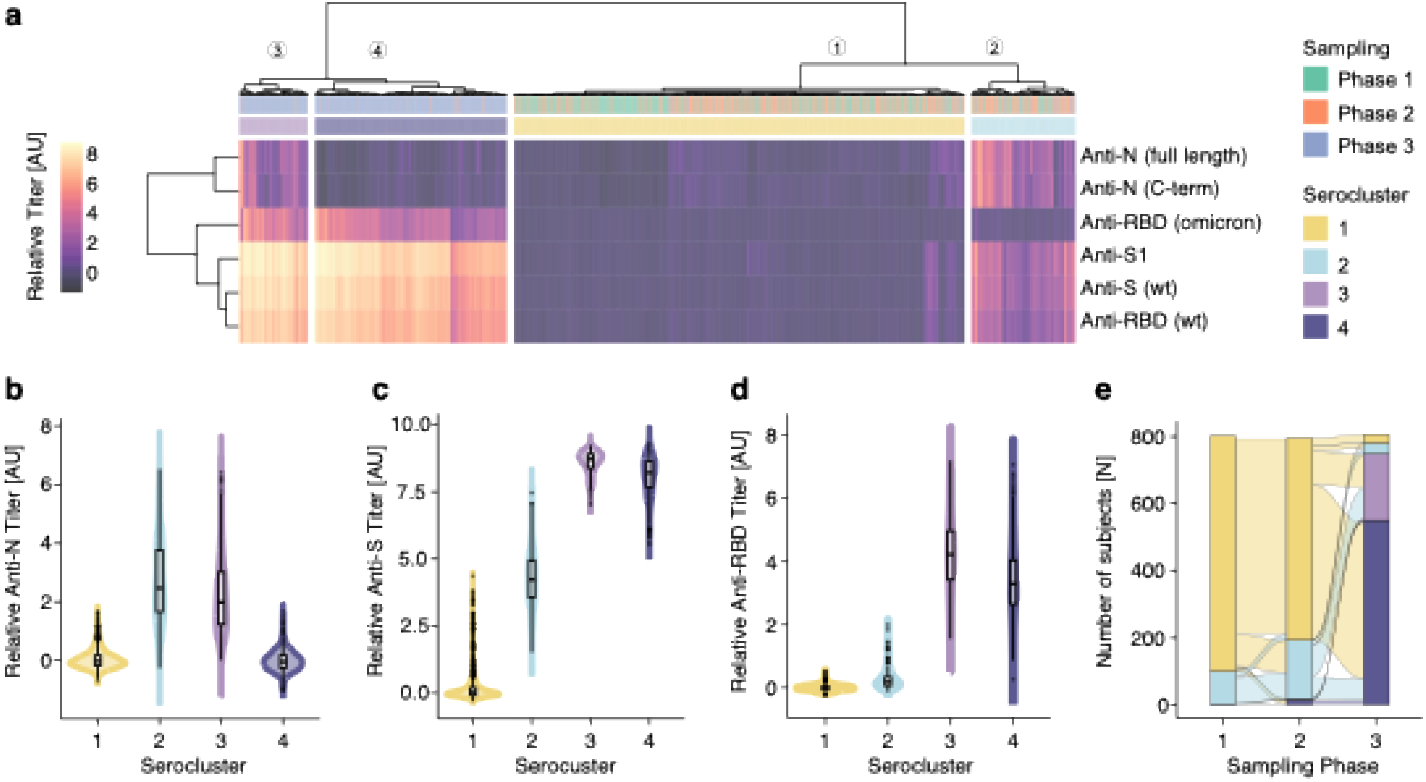
Serological analysis. (a) The heatmap shows the cluster analysis of multi-analyte serology assays (2x N, 2x RBD, 2x S proteins) to define four distinct seroclusters: SC1 contained 1220 samples from 722 donors who were deemed seronegative at the timepoint of sampling. SC2 contained 306 samples from 211 donors who were deemed infected. SC3 contained 203 samples from 199 donors who were deemed infected and vaccinated. SC4 contained 565 samples from 552 donors who were deemed vaccinated only (b-d). Serological profiles per serocluster (e) Distribution of seroclustes per sampling phase.

Serocluster 1 samples were collected mostly during Phase 1 and Phase 2 (Fig. 2e), hence prior to the start of vaccinations of the general population in Sweden. Within the phases, there were no differences in age, sex or smoking between the seroclusters (Tab. 2). Among the 99 S+ subjects in Serocluster 2 from Phase 1, 84.9% reported symptoms of suspected or confirmed COVID-19 since February 2020 as compared to 38.5% in Serocluster 1. Further, there were significantly higher proportions of subjects reporting a positive antibody or PCR test (43.3%) and having a confirmed infection defined as a SmiNet-case (11.1%) in Serocluster 2 compared to Serocluster 1. A similar pattern was observed in Phase 2, where Serocluster 2 had the highest proportions of individuals with self-reported PCR or antibody test (42.9%) and SmiNet-confirmed infection (38.9%) compared to the other clusters (Tab. 2).

**Table 2.** Serocluster demographics.

|  | Phase 1 (n=802) |  |  |  |  | Phase 2 (n=796) |  |  |  |  | Phase 3 (n=806) |  |  |  |  |
| --- | --- | --- | --- | --- | --- | --- | --- | --- | --- | --- | --- | --- | --- | --- | --- |
| Serocluster | 1 | 2 | 3 | 4 | p | 1 | 2 | 3 | 4 | p | 1 | 2 | 3 | 4 | p |
| n | 702 | 99 | 1 | 0 |  | 603 | 175 | 3 | 15 |  | 25 | 32 | 199 | 550 |  |
| Female (yes) | 455 (64.8) | 57 (57.6) | - | - | 0.18 | 387 (64.2) | 110 (62.9) | - | 13 (86.7) | 0.19 | 14 (56.0) | 20 (62.5) | 130 (65.3) | 351 (63.8) | 0.82 |
| Age (years) | 25.4 (23.7-26.7) | 25.3 (23.7-26.5) | (23.9-27.2) | - | 0.49 | 25.9 (24.0-27.2) | 25.7 (24.2-27.3) | - | 26.4 (25.4-27.6) | 0.44 | 27 (25.0-27.6) | 26.7 (24.9-27.7) | 26.5 (24.9-27.8) | 26.6 (24.9-27.8) | 0.20 |
| Smoking (yes)<sup>1</sup> | 91 (13.0) | 20 (20.2) | - | - | 0.06 | 64 (10.6) | 24 (13.7) | - | 0 (0.0) | 0.23 | 2 (8.0) | 8 (25.0) | 19 (9.7) | 51 (9.3) | 0.06 |
| Symptoms (Since February 2020) | 269 (38.5) | 84 (84.9) | - | - | <0.001 | - | - | - | - | - | - | - | - | - | - |
| Symptoms (Since August 2020) | - | - | - | - |  | 247 (41.0) | 99 (56.6) | - | 89 9 (53.3.) | 0.001 | - | - | - | - | - |
| Symptoms ≥ 2 months<sup>1</sup> | 10/268 (3.7) | 2/84 (2.4) | - | - | 0.74 | 11/247 (4.5) | 4/99 (4.0) | - | 1/8 (12.5) | 0.49 | 1/10 (10.0) | 3/20 (15.0) | 26/163 (16.0) | 27/207 (13.0) | 0.87 |
| Symptoms ≥ 3 months<sup>1</sup> | - | - | - | - |  | - | - | - | - | - | 1/10 (10.0) | 2/20 (10.0) | 21/163 (12.9) | 25/207 (12.1) | 0.99 |
| Self-reported positive antibody-test | 3 (0.4) | 35 (35.4) | - | - | <0.001 | 3 (0.5) | 25 (14.3) | - | 1 (6.7) | <0.001 | 3 (12.0) | 19 (59.4) | 106 (53.8) | 103 (18.8) | <0.001 |
| Self-reported positive PCR-test | 1 (0.1) | 10 (10.1) | - | - | <0.001 | 6 (1.0) | 57 (32.6) | - | 0 (0.0) | <0.001 | 0 (0.0) | 6 (18.8) | 97 (49.2) | 41 (7.5) | <0.001 |
| Self-reported positive PCR- or antibody-test | 4 (0.6) | 43 (43.3) | - | - | <0.001 | 9 (1.5) | 75 (42.9) | - | 1 (6.7) | <0.001 | 3 (12.0) | 21 (65.6) | 156 (78.4) | 108 (19.6) | <0.001 |
| SmiNet-case (yes)<sup>2</sup> | 0 (0.0) | 11 (11.1) | - | - | <0.001 | 7 (1.2) | 68 (38.9) | - | 0 (0.0) | <0.001 | 0 (0.0) | 9 (28.1) | 112 (56.3) | 31 (5.7) | <0.001 |
| Days since SmiNet-case | ND | 67 (35-144) | - | - |  | 157 (70-272) | 74 (10-252) | - | - | 0.015 | NA | 257 (11-415) | 242 (1-536) | 373 (6-621) | <0.001 |
| New SmiNet-case<sup>3</sup> | - | - | - | - |  | 5 (0.8) | 60 (34.3) | - | 0 (0.0) | <0.001 | 0 (0.0) | 3 (9.4) | 66 (33.2) | 5 (0.9) | <0.001 |
| Vaccinated COVID-19<sup>4</sup> | 0 (0.0) | 0 (0.0) | - | - |  | 3 (0.5) | 8 (4.6) | - | 12 (80.0) | <0.001 | 1 (4.0) | 14 (43.8) | 195 (98.0) | 541 (98.7) | <0.001 |
| Not vaccinated | - | - | - | - |  | 600 (99.5) | 167 (95.4) | - | 3 (20.0) | <0.001 | 24 (96.0) | 18 (56.3) | 4 (2.0) | 7 (1.3) | <0.001 |
| 1 vaccine dose only | - | - | - | - |  | 3 (0.5) | 7 (4.00) | - | 2 (13.3) | - | 0 (0.0) | 1 (3.1) | 8 (4.0) | 8 (1.5) | - |
| 2 vaccine doses only | - | - | - | - |  | 0 (0.0) | 1 (0.6) | - | 10 (66.7) | - | 1 (4.0) | 13 (40.6) | 176 (88.4) | 516 (94.2) | - |
| 3 vaccine doses | - | - | - | - |  | 0 (0.0) | 0 (0.0) | - | 0 (0.0) | - | 0 (0.0) | 0 (0.0) | 11 (5.5) | 17 (3.1) | - |
| Days since vaccine dose 1 | - | - | - | - |  | 8 (4-9) | 21 (11-106) | - | 45 (11-111) | 0.011 | 123 | 247 (43-290) | 133 (24-401) | 121 (12-352) | <0.001 |
| Days since vaccine dose 2 | - | - | - | - |  | - | 77 | - | 13 (6-34) | 0.11 | 83 | 179 (1-237) | 89 (7-251) | 78 (4-305) | <0.001 |
| Days since vaccine dose 3 | - | - | - | - |  | - | - | - | - | - | - | - | 21 (3-62) | 21 (1-67) | 0.91 |
| Hospital SARS-CoV-2 serology test positive<sup>5</sup> | - | - | - | - |  | 33 (5.5) | 167 (95.4) | - | 14 (93.3) | <0.001 | - | - | - | - | - |

Continuous variables were analysed by the Mann-Whitney U-test or Kruskal-Wallis test and are displayed as median with range. Dichotomous variables were analysed by the Fisher's exact test and are displayed as number and percentage.<sup>1</sup>Self-reported in Phase 1-3 questionnaires; <sup>2</sup>SmiNet (a national register for notifiable transmittable diseases in Sweden, covering all positive PCR tests (Public Health Agency of Sweden. SmiNet. In Swedish. Available at: https://www.folkhalsomyndigheten.se/sminet)). A SmiNet-case was defined as a positive entry date in SmiNet preceding the date of DBS filter paper sampling in each respective phase; <sup>3</sup>SmiNet-case in the current phase, but not in the preceding phase; <sup>4</sup>Information on vaccination against COVID-19 was obtained by linkage to the National Vaccination Register. COVID-19 vaccination was defined as receiving the vaccination prior to the date of DBS filter paper sampling in each respective phase. <sup>5</sup>Determined from Karolinska University Hospital Laboratory standard SARS-CoV-2 IgG serology test performed at the clinical examination in Phase 2

In Phase 2 and 3, Serocluster 4 had the highest proportion of vaccinated individuals (Phase 2 80%, Phase 3 98.7%). In Phase 3, this Serocluster also had the shortest median time between the second vaccine dose and sample collection (78 days). Serocluster 4 contained most samples from Phase 3 (Fig. 2e), which is consistent with the vaccination rollout (Fig. 1d). In Phase 3, Serocluster 3 likely constituted a vaccinated group that had been previously infected, based on the high prevalences of self-reported PCR- and antibody tests. Similarly, Serocluster 2 in Phase 3 might represent vaccinated and previously infected subjects, for which vaccinations were given furthest back in time, and there were also fewer new SmiNet-cases in this cluster (Tab. 2). Of all subjects, only 25 (~3%) remained S- and N-during all sampling periods, as represented by Serocluster 1 in Phase 3. The majority of these 25 subjects also belonged to Serocluster 1 in Phase 2. The effect of time and possible waning of antibody levels is further described in the supplementary.

In summary, this confirmed the expected seroprevalence rates over time, adding a refined classification from self-sampling for time-resolved immune states.

### Identification of auto-reactive antibodies (AAbs) against interferons

Next, we measured AAbs against 21 interferons (IFNs) to gain further insights into pre-existing or newly developed immune states. The proportions of AAb+ subjects within each phase are displayed in Tab. 3 and Fig. 3a. In this narrow-aged study, the individual anti-IFN AAbs were detected at low frequencies, with median prevalences of 3.5%, 3.0% ad 3.2% in Phases 1-3, respectively. The highest number of AAb+ individuals were found for Interferon Gamma Receptor 2 (IFNGR2) with 8.2%, 9.2% and 10.4% in Phases 1-3, and Interferon alpha/beta receptor 1 (IFNAR1) with 6.6%, 8.5%, and 10.9%. For most IFNs, the percentage of AAb+ subjects remained stable across phases, except for IFNAR1 and IFNAR2, which increased (Tab. 3, Fig. 3a). There was a high intra individual correlation between the anti-IFNA and anti-IFNL AAbs (Fig. S6). We then divided the subjects based on their longitudinal AAb pattern: being seropositive across all phases (persistent AAb+), being seronegative across all phases (always AAb-) and all remaining patterns across phases (changing) (Tab. 3). For eight AAbs, we observed ≥10 AAb+ subjects in all phases, which we included to investigate sex, and COVID-19 symptoms, infection, serocluster belonging and vaccination status. We found a higher proportion of females in the changing group for IFNL3, and lower proportions of females in the changing and always positive groups for IFNA8 and IFNG (Fig. 3b, Tab. S1). We did not find any associations of AAb patterns with infection, serocluster belonging or vaccination. However, we observed a higher percentage of persistent AAb+ subjects reporting COVID-19-related symptoms lasting for ≥ 2 months and ≥ 3 months within the groups being always positive for IFNGR2 and IFNAR1 (Figs. 3c and 3d, Tab. S1).

**Table 3.**
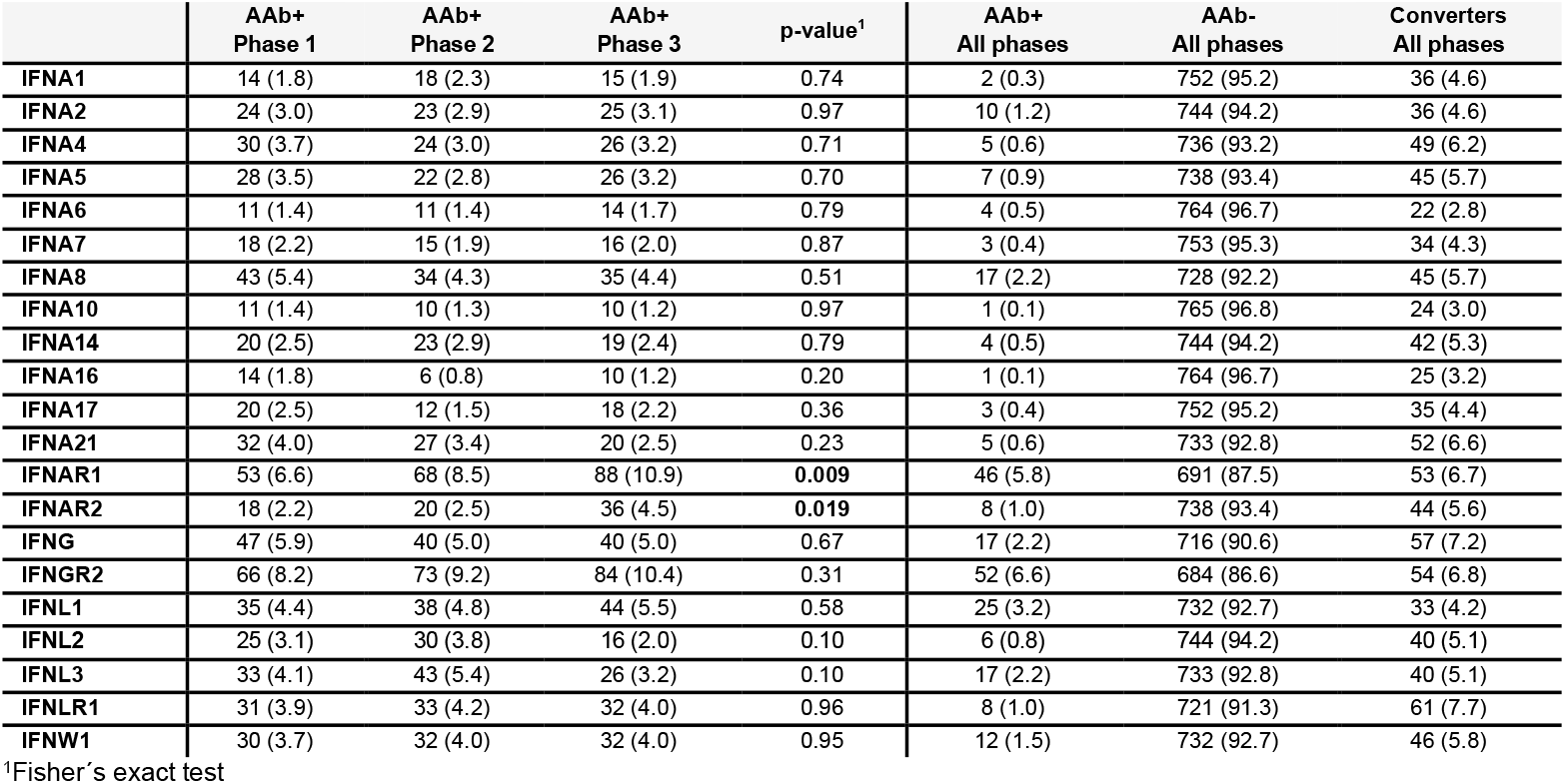
Anti-interferon AAb positivity rates per and across sampling phases.

**Fig. 3:**
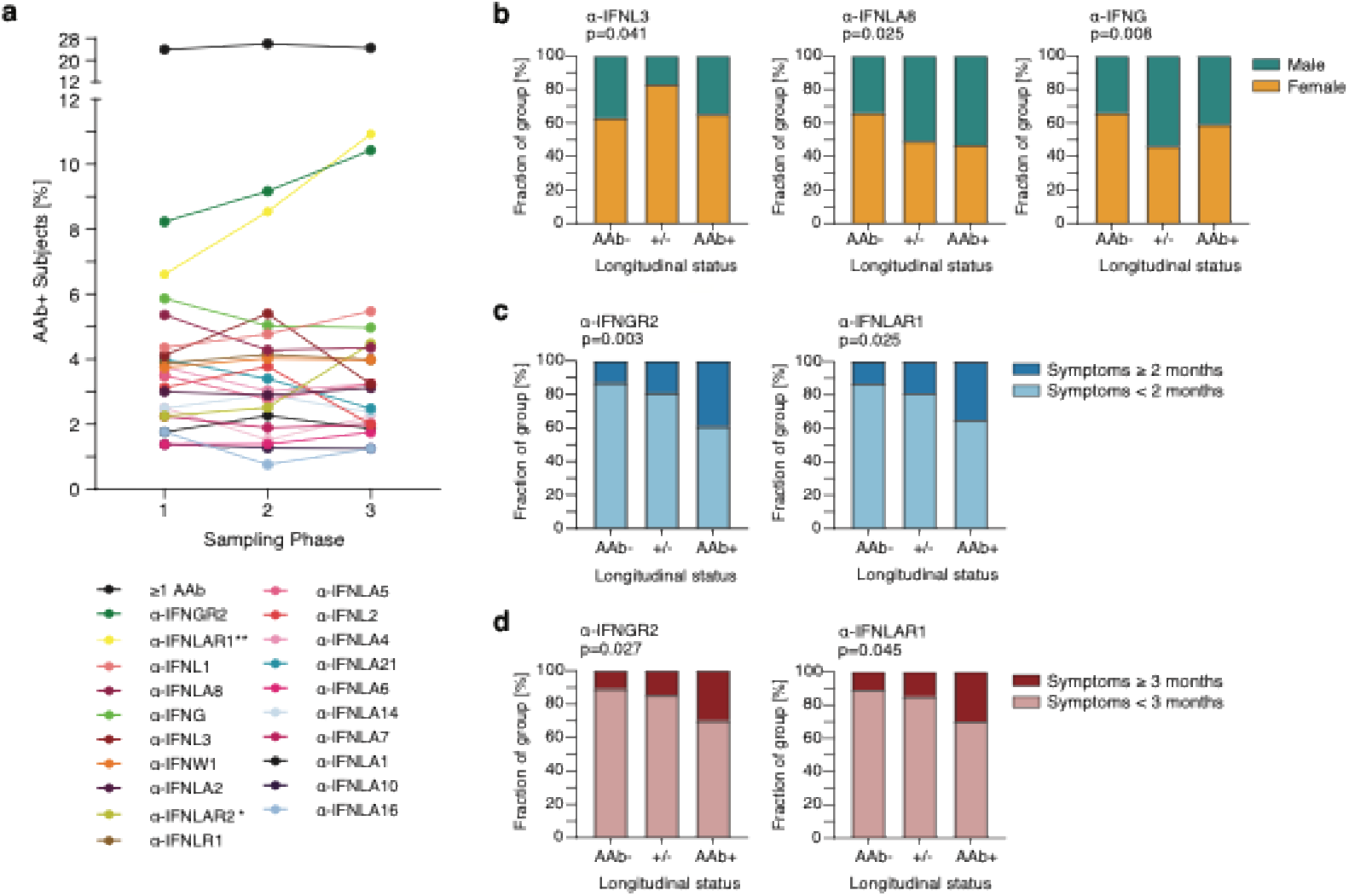
Autoantibody analysis. (a)The proportion of subjects seropositive for ≥1 AAb or each measured AAb within each Phase 1 to 3. (b) The proportion of males and females within the groups being always negative, always positive and changing in Phase 1 to 3 for anti-IFNL3, anti-IFNA8 and anti-IFNG. (c,d) The proportion of subjects with COVID-19-related symptoms ≥2 months (c) and ≥3 months (d) within the groups being always negative, always positive and changing in Phase 1 to 3 for anti-IFNGR2 and anti-IFNAR1. Data was analyzed by the Fisher’s exact test.

The longitudinal anti-IFN AAbs patterns revealed a low frequency prevalence of pre-existing and persistent AAbs in a young adult population and highlighted a possible link between anti-IFNGR2 and anti-IFNAR1 AAbs and extended COVID-19-related symptoms.

### Global analysis of the circulating DBS proteins

To expand the molecular phenotypes, we measured 365 inflammation-related proteins in the same DBS, applying protocols developed during previous population studies^16,17^.

First, we performed principal component analysis (PCA) on all proteins (n = 365) in all samples that passed QC controls (n = 2394) to evaluate the higher dimensional data. We observed samples to group by time point (Fig. S7a), indicating the overall impact of SARS-CoV-2 exposures on the circulating DBS proteome. Investigating principal components (PCs) 1-3 with clinical variables revealed that PC1 correlated with sampling phase (Rho = 0.76), sampling date (Rho = 0.75), number of vaccine doses (Rho = 0.75), serocluster (Rho = 0.71), and levels of anti-S Abs (Rho = 0.65) (Fig. S7c). Furthermore, the PC1 correlated with mean protein levels for the sample (Rho = 0.95), confirming the impact of pre-analytical and clinical factors. Out of the 365 proteins, 89 (24.4%) had a PC1-correlation beyond rho = ± 0.5 (Fig. S7b,d). On the other hand, the available clinical variables or design features did not correlate with PC2 and PC3. For the proteins, IL10 levels were negatively correlated with PC2 (Rho = −0.73). For PC3, there were 10 proteins with a correlation beyond Rho < −0.6, and a positive correlation to IL10 of Rho = 0.54 (Fig. S7d).

### Associations of circulating DBS proteins with clinical traits

Based on our initial assessment, we investigated proteomic associations with the demographic factors age, sex, BMI and smoking in Phase 2 when a clinical examination follow-up was conducted. We found 133 proteins (36%) being significantly associated with BMI (Fig. 4a). Secretoglobin family 3A member 2 (SCGB3A2) and the WAP, follistatin/kazal, immunoglobulin, kunitz and netrin domain containing 2 (WFIKKN2), and Advanced glycosylation end-product specific receptor (AGER) were top ranked proteins. Compared to previous population-wide studies, the presented cohort was more homogenous and narrow in age, between 23.9 and 27.2 years, hence only five proteins were found to be significantly associated. This included Nuclear distribution C, dynein complex regulator (NUDC), Src kinase associated phosphoprotein 2 (SKAP2) and Phosphoinositide-3-kinase adaptor protein 1 (PIK3AP1) as the top ranked. For sex, we found a total of 198 (51%) significantly associated proteins, with Peroxiredoxin-5, mitochondrial (PRDX5), Triggering receptor expressed on myeloid cells 2 (TREM2) and Fission, mitochondrial 1 (FIS1) being the three most significant ones (Fig. 4b). Compared to UK Biobank (UKB), we did not find any proteins associated with smoking in our cohort, which is likely due to the relatively lower number of smokers (~12% in all three phases) and shorter smoking periods. The results, summarized in Figs S8-9, were compared to the betas and p-values from a DBS survey^16^ and the large scale UKB plasma study^1^. For BMI and sex Many proteins-trait associations were consistent between BAMSE and the UKB data (Fig. S8), even though the BAMSE cohort consists of a narrow aged younger population and DBS instead of plasma sample. Associations of protein levels with sex were also concordant between BAMSE and a previous DBS study^16^ (Fig. S9).

**Fig. 4:**
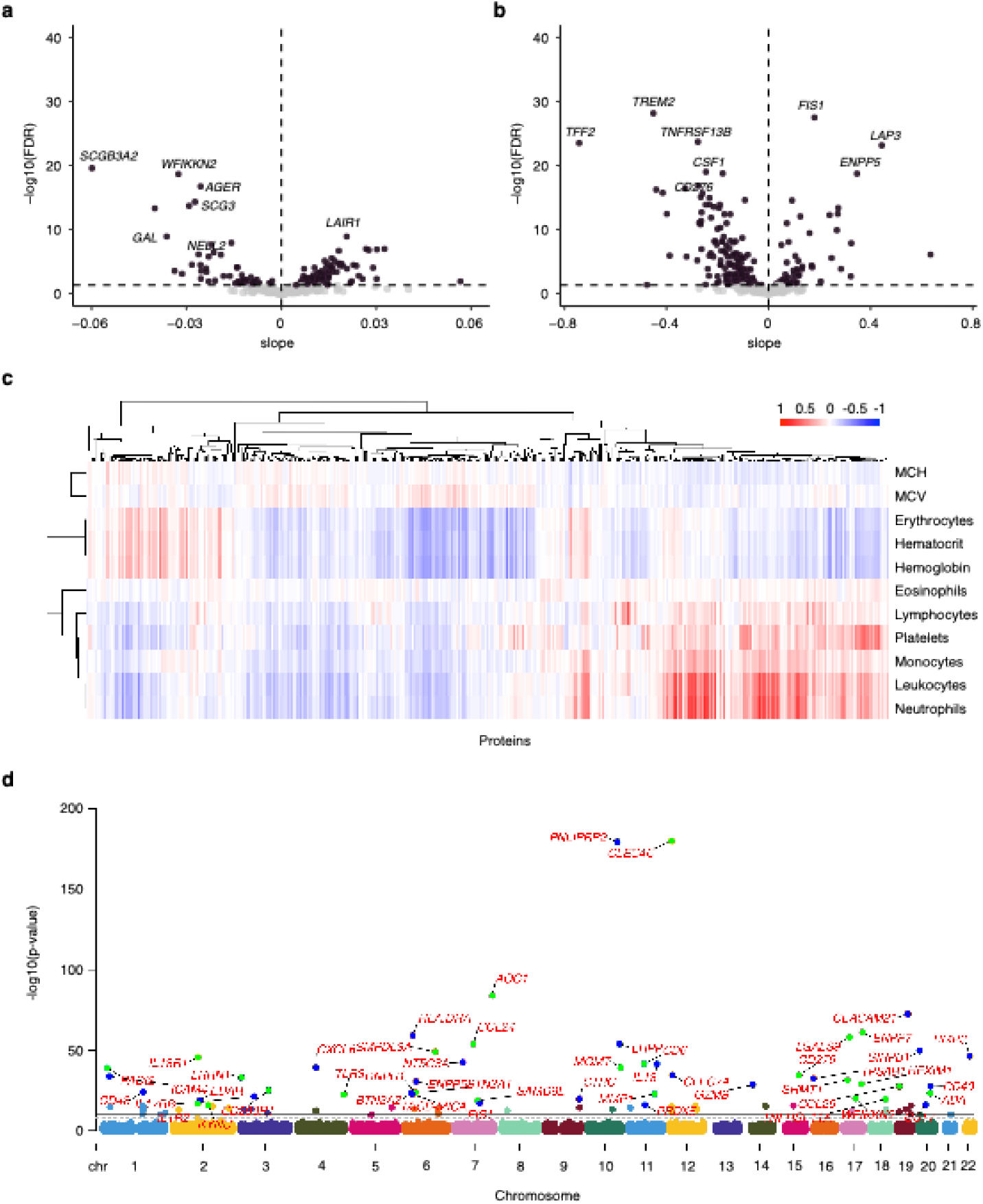
Proteomic analysis. (a, b) Linear associations between protein levels and (a) BMI and (b) sex. The slope is plotted against the −log10(FDR) value. Purple dots mark significant proteins. Top 10 significant protein names are stated in the plot. (c) Spearman correlations between complete blood count (CBC) with differential parameters and 365 proteins at Phase 2 of the COVID-19 follow-up (n=782). Cell color indicates degree and directon of Spearman’s rho correlation. Dendrograms reflect hierarchical clustering using average linkage on distance matrices. (d) The Manhattan plot shows the top 50 most significant pQTLs.

### Investigation of circulating DBS proteins for pre-analytical factors

To evaluate the impact of time between sampling and analysis for stored DBS, we studied protein levels of 25 persistently seronegative donors (all Phases). We identified 99 of 365 (27%) proteins to increase or decrease (Bonferroni p < 0.01) with prolonged storage (Fig. S10). We assessed enrichment of storage-sensitive proteins across protein classes and found intracellular proteins enriched among storage-sensitive ones (FDR < 0.01), suggesting a higher susceptibility to degradation compared to other classes. This analysis revealed the profound impact of the exposure and sensitivity of blood proteins to pre-analytical design aspects.

### Relationship between complete blood cell counts and circulating DBS proteins

To explore how blood cells in DBS contribute to the data, we studied the distribution of complete blood cells (CBC) and associations with levels of DBS proteins.

We used 782 samples from Phase 2 (Fig. S11), of which the majority had blood counts (Tab. 4) within the reference ranges provided by the clinical laboratory^18,19^. Notably, leukocytes, neutrophils, and platelets were positively correlated (rho > 0.5) with several proteins, including PLAUR, NCF2, OSM, LAT, and CXCL3, while erythrocytes, hematocrit, and hemoglobin showed fewer and predominantly negative protein correlations, as shown in Fig. 4c. Hierarchical clustering revealed clusters of cell types, with protein correlations overlapping across CBC parameters shown in Fig. S12. Leukocytes and neutrophils accounted for most of the FDR-significant associations, whereas eosinophils, mean corpuscular hemoglobin (MCH), and mean corpuscular volume (MCV) had relatively few FDR-significant correlations beyond rho = ±0.5. Fig. S13 displays the strongest significant positive and negative correlations for each CBC parameter.

**Table 4.** Blood cell counts of Phase 2 seroclusters.

| Phase 2 samples | All | Serocluster 1 | Serocluster 2 | Serocluster 4 | p-value |
| --- | --- | --- | --- | --- | --- |
| n | 799 | 595 | 174 | 15 | - |
| Basophils (x10<sup>9</sup>/L) | 0.0 (0.0-0.1) | 0.0 (0.0-0.1) | 0.0 (0.0-0.1) | 0.0 (0.0-0.0) | 0.99 |
| Eosinophils (x10<sup>9</sup>/L) | 0.1 (0.0-1.0) | 0.1 (0.0-1.0) | 0.1 (0.0-0.8) | 0.1 (0.0-0.7) | 0.18 |
| Leucocytes (x10<sup>9</sup>/L) | 5.8 (3.0-11.4) | 5.8 (3.0-11.4) | 5.6 (3.1-9.8) | 6.6 (3.7-9.1) | 0.20 |
| Lymphocytes (x10<sup>9</sup>/L) | 1.8 (0.7-3.8) | 1.8 (0.7-3.8) | 1.9 (0.7-3.6) | 1.9 (1.5-2.5) | 0.46 |
| Monocytes (x10<sup>9</sup>/L) | 0.5 (0.2-1.5) | 0.5 (0.2-1.0) | 0.5 (0.2-1.5) | 0.4 (0.3-0.6) | 0.61 |
| Neutrophils (x10<sup>9</sup>/L) | 3.2 (0.9-8.8) | 3.2 (0.9-8.8) | 2.9 (1.3-7.0) | 3.7 (1.7-7.0) | 0.05 |
| Erythrocytes (x10<sup>12</sup>/L) | 4.6 (3.5-6.2) | 4.6 (3.6-6.1) | 4.6 (3.5-5.8) | 4.6 (3.8-6.2) | 0.85 |
| EVF | 0.4 (0.3-0.5) | 0.4 (0.3-0.5) | 0.4 (0.3-0.5) | 0.4 (0.4-0.5) | 0.41 |
| Hemoglobin (g/L) | 138 (77-179) | 138 (77-179) | 139 (103-164) | 134 (120-174) | 0.25 |
| Thrombocytes (x10<sup>9</sup>/L) | 253 (88-514) | 253 (88-514) | 253 (150-453) | 287 (163-442) | 0.15 |
| MCV (fL) | 90 (68-105) | 90 (68-105) | 90 (81-102) | 90 (83-97) | 0.09 |
| MCH (pg) | 30 (19-35) | 30 (19-35) | 30 (25-34) | 30 (27-32) | 0.12 |
| HbA1c mmol/mol | 31 (20-48) | 31 (24-39) | 31 (25-41) | 32 (27-48) | 0.80 |

Data was analysed by the Kruskal-Wallis test and is displayed as median with range. EVF: erythrocyte volume fraction/hematocrite (percent of blood volume). MCV: erythrocyte mean corpuscular volume. MCH: mean corpuscular hemoglobin.

Next, we employed linear regression models to investigate associations between DBS proteins and CBC parameters, revealing the largest estimates for hematocrit and proteins TFF2, FST, and ANGPTL4. The highest number of protein associations were obtained for leukocytes and neutrophils (215 and 212 proteins, respectively; Fig. S14). Similar associations were observed in the adjusted models (using alone sex or sex, smoking, and BMI). Again, the largest estimates were detected between hematocrit and TFF2, MICB/MICA, and FST, and the highest number of protein associations for leukocytes and neutrophils.

To check the consistency of CBC-protein associations, we evaluated inter-correlations for proteins previously measured in plasma^20^ and now in DBS (Fig. S15). With 51 overlapping proteins, strong inter-correlations were observed, including MMP1, IL12B, IL10RB, and CCL4. Finally, we contrasted the CBC~plasma and CBC~DBS protein correlations (Fig. S16). We found directionally concordant associations for white blood cell (WBC) parameters (leukocytes and neutrophils), while red blood cell (RBC) parameters (erythrocytes, hematocrit, and hemoglobin) displayed inverse correlations between DBS and plasma.

In summary, we determine the links between dried (DBS) and liquid (plasma) proteins to CBC and found these to be most notable for leukocytes and neutrophils. Expectedly, the presence of RBCs in DBS has a different impact than plasma samples.

### Contribution of genetic variation on the levels of circulating DBS proteins

To map the genetic determinants of inflammation protein expression, we performed a genome-wide protein quantitative trait locus (pQTL) analysis, including 650 subjects for whom both DBS proteomic and imputed genotype data were available.

We identified a total of 664 significant pQTLs spanning 65 proteins with genome-wide significance (p < 1.3×10^−10^). We resolved a clear partitioning of regulatory mechanisms: 384 (58%) of these associations were local (*cis*-pQTLs, defined as variants within ±1 Mb of the gene encoding the associated protein), while 280 (42%) acted distantly (*trans*-pQTLs) (Fig. 4d). This agreed with the previous studies^1,21^ and highlights the quality of the DBS proteome data also in relation to pQTL analyses, as the *cis*-pQTLs could be validated. Among the identified pQTLs, all the *cis*-pQTLs validated against the previously reported ones in UKB-PPP, SCALLOP or OpenGWAS resources and 135 *trans*-pQTLs were regarded as novel at the time of investigation.

### Facets of longitudinal changes of circulating DBS proteins

With insights from the impact of exposure, design, clinical traits, blood cell counts, and association to genetics, we evaluated changes across the three consecutive sampling phases.

To assess the facets of longitudinal protein variability, inter-phase correlations and intra-class correlation coefficients (ICCs) were calculated (Fig. S17a,b). Among all 365 proteins, 18 (4.9%) were stable (ICC ≥ 0.75) between Phase 1~2, Phase 2~3, or Phase 1~3, and 14 (3.8%) were stable in all. Notably, the PNLIPRP2, MMP1, CLEC4C, TPSAB1 and

ICAM4 were among the most stable proteins across all comparisons (ICC > 0.83) (Fig. S17c-g). Reassuringly, all of these had *cis*-pQTLs (MMP1, CLEC4C, TPSAB1), or *trans*-pQTLs (PNLIPRP2, ICAM4) at genome-wide significance. A protein with stable profile between all phases without a pQTLs was Galanin peptides (GAL).

In addition to protein-level investigations, we studied how the profile of the 800 donors changed over time. Performing paired subject correlations between phases, mean Spearman correlations where higher in matched pairs (mean rho = 0.25) compared to random pairs (mean rho = 0.02) (Fig. S17k). We observed the strongest matched pair correlations between lower exposure sampling of Phase 1~2 compared to Phase 1~3, relating to the vaccine rollout and Omicron wave. However, this also applied when limiting persistently seronegative individuals from Serocluster 1 (Fig. S17l), pointing at the possible impact of pre-analytical factors.

### Capturing the sampling time-dependent dynamic of circulating DBS proteins

Lastly, we investigated time in relation to pandemic exposures and DBS protein levels. To minimize the impact of imbalanced sample sizes, we adjusted the number of donors between groups using random resampling for larger groups.

When comparing protein level changes between the I– and I+ subjects, we found leucine aminopeptidase 3 (LAP3) to be consistently elevated in DBS samples close to an infection (median FDR = 0.00005) (Figs. S18a). In cross-cluster comparisons within each phase, LAP3 remained significantly elevated in the infected group. Applying bootstrapping for Phase 2 group comparisons, LAP3 remained elevated in all iterations (median FDR = 5×10^−6^, Fig. S18b). The previously COVID-associated protein Galectin-9 (LGALS9) was significantly elevated and selected in 74% of the iterations (median FDR = 0.006, Fig. S18b). No significant differences were observed between previously infected subjects and the seronegative group (Fig. S19a). When comparing previously infected subjects to more recently infected subjects, LAP3 and LGALS9 were significantly elevated in the newly infected group, while TRAF2 was lower (Figs. S19b,e-g).

To further investigate the timing effect of when infection and vaccination occurred in relation to DBS self-sampling, we compared longitudinal changes in selected subjects of Phase 2 and 3. We compared 92 seronegative subjects from Phase 2 who were vaccinated and infected in Phase 3 (I–V– converting to I+V+) to 92 infected from Phase 2 who received the vaccine before sampling in Phase 3 (I+V– converting to I+V+). For Phase 2, LAP3 was significantly elevated in the infected versus seronegative group, and for Phase 3 LAP3 was elevated in the more recently infected group (Fig. S19c). There was also a significant difference in longitudinal LAP3 level changes between these groups, indicating a potential role of LAP3 in response to infection. There were no significant differences in LAP3 changes when comparing I–V+ individuals with I+V+ subjects showing waning anti-N antibodies (Fig. S19d).

Next, we transformed the protein levels into z-scores to better compare the number of samples changing along the time axis of matched pre- and post-infection or vaccination samples (Figs. 5a-c). As shown in Fig. 5, several proteins showed significant shift in number of samples beyond a z-score threshold (z < –2 or z > 2). After infection, significantly more samples had LAP3 z-score values > 2 (FDR = 0.0026, Fig. 5d) for about a month since PCR diagnosis (mean 29.5 days, median 27.5 days). Dissecting the data into a 30-day interval around infection, LAP3 levels were significantly altered (FDR = 0.0005) between pre- and post-infection among PCR-confirmed subjects (Figs. S18c-d) before declining. Only three subjects collected samples within 10 days of PCR confirmed infection, limiting the resolution to further study of even shorter temporal changes. Notably, LAP3 elevation was not observed when comparing protein levels pre and post vaccination (Fig. S18e).

**Fig. 5:**
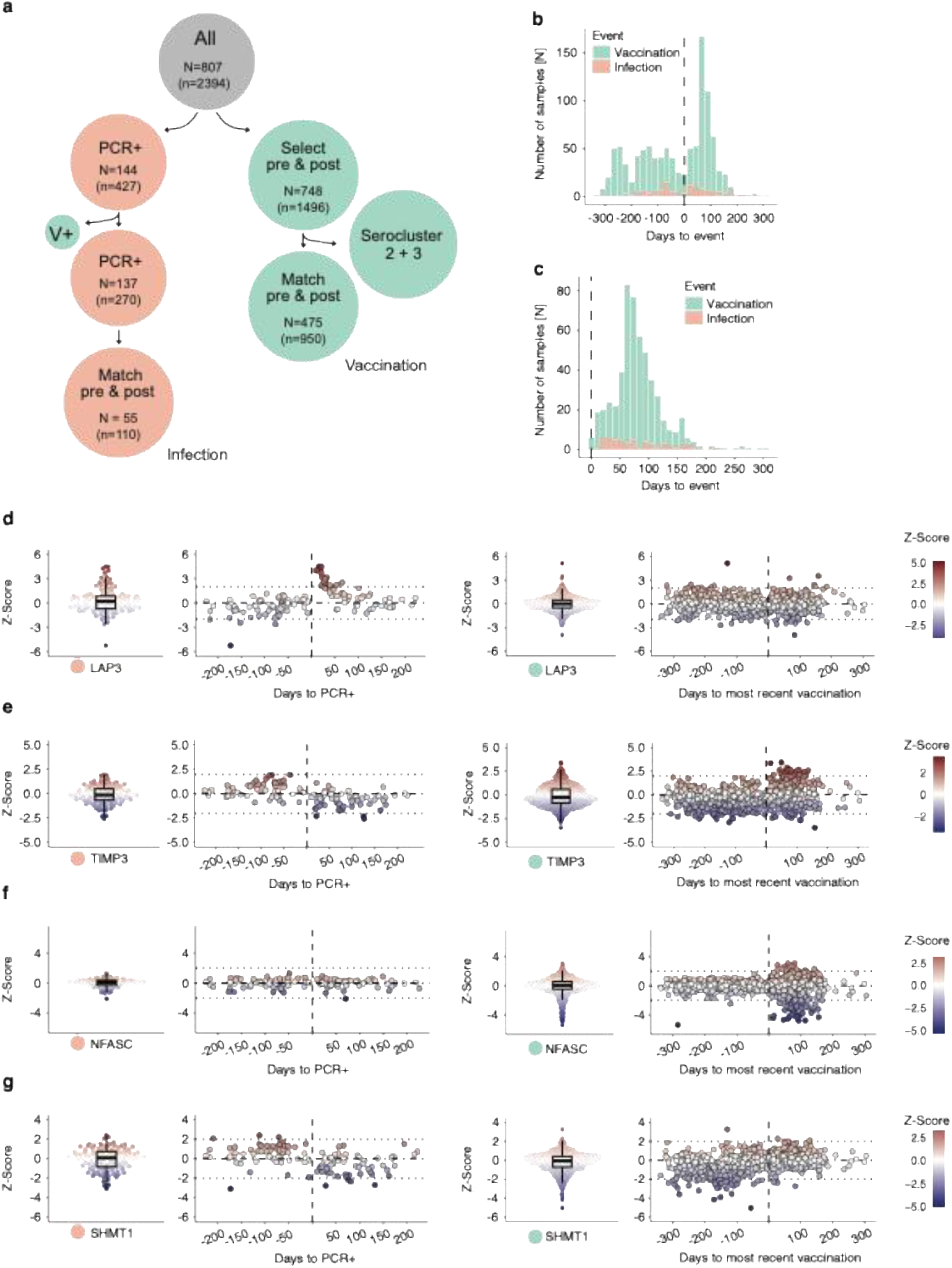
Proteins in relation to infection and vaccination. (a) Overview of selection of infected and vaccinated samples. (b) Histogram of time since event with closest vaccination dose in green and PCR-confirmed infection in orange for with samples pre and post infection. (c) Histogram shows only samples post infection. (d-g) Protein z-scores plotted against days since PCR-confirmed infection (left panel) and closest vaccination dose (right panel). The boxplots show the 25% and 75% quartiles with the median in the center.

Following vaccination, 20 proteins had significantly more samples above the z-score threshold. Among these, TIMP metallopeptidase inhibitor 3 (TIMP3, Fig. 5e) and Phospholysine phosphohistidine inorganic pyrophosphate phosphatase (LHPP), and SRSF protein kinase 2 (SRPK2) were the most significant. In contrast, two proteins had significantly more samples before vaccination. For values below the lower z-score threshold, four proteins had significantly more samples after vaccination, with Neurofascin (NFASC, Fig. 5f) being most significant. Conversely, 54 proteins showed significantly more samples below z-score threshold before vaccination. Serine hydroxymethyltransferase 1 (SHMT1, Fig. 5g) was most significant with 41 out of 475 samples before compared to 3 samples after vaccination.

The identification of transient and exposure-dependent alterations in self-sampled DBS adds a new dimension to studying dynamic molecular phenotypes.

## Discussion

Comprehensive molecular analyses of self-sampled DBS elucidated molecular trajectories of the immune response against natural infection and vaccination. Being among the first, the study demonstrates the suitability of at-home self-sampling as an alternative to conventional clinical sampling for repeated and robust multi-molecular monitoring.

Multi-analyte serology revealed the expected distinction between infections (N+, S+ and RBD+) or vaccination (N–, S+ and RBD+), adding to donor-provided and being consistent with previous reports^16^. Our IgG data is further backed by PCR-confirmed infection dates and vaccination records, and repeated sampling improving the resolution, stratification and estimations of antibody decay. By using data-driven seroclustering, we improved the sample stratification according to their actual humoral antibody responses at the time of sampling, rather than relying solely on self-reported data.

In addition to anti-SARS-CoV-2 Abs, we detected auto-Abs (AAbs) against several IFN proteins in the same assay. Despite inconsistencies across different anti-IFN autoimmunity reports^22^, the longitudinal design enabled us to differentiate between occasional and consistent presence of anti-IFN AAbs. This revealed clinical phenotypes associated with pre-existing and persistent AAbs, rather than new-onset AAbs, supporting the necessity of considering longer trajectories of autoimmune profiles in health assessment exercises. The differences to previous applications of the assay in a self-sampling survey of the general population can be explained by the wider age range and cohort heterogeneity ^16^.

DBS offers a minimally invasive sample compatible with high-resolution multi-molecular analyses^10^. Still, the impact of the cells, such as RBCs or WBCs, over the soluble and secreted components of the plasma proteome^23^ remains unclear, because upon drying, the cells leak additional proteins into the DBS. While this offers opportunities to identify previously inaccessible proteins, leakage also poses a risk of adding possible confounders to the analyses. For example, our data suggests that secreted and membrane proteins, possibly due to glycosylation, appear to be more stable during storage time than intracellular proteins. Using the clinical cell count data, we extracted the missing links between cell count and protein abundance, lifting the impact of how, for example, hematocrit or platelet counts shape the protein profiles. This enables a more informed mechanistic interpretation of protein abundance and origin and supports an interchangeable conversion of clinically established and advanced omics measurements.

Many of the protein associations with BMI and sex identified were consistent in direction and magnitude between BAMSE DBS data and UKB plasma data^1^, supporting the utility of and validity of the presented data. The lack of common age-associated proteins with UKB plasma data or our previous general population DBS study^16^ is explained by the narrow age range of our participants (± 4 years vs. 50+ years). This mismatch also drove inconsistencies in other age-related traits, such as for the duration of smoking in years.

We also conducted GWAS analysis with the DBS proteomes data. Aligned with previous reports on plasma^1,24,25^, we found pQTLs for many targets, including MMP1, PNLIPRP2, and TPSAB1. This supports the notion that we observed intended targets in our DBS assays, as recently suggested in a study on newborns^21^. This is particularly interesting as many inflammatory proteins often vary due to exposure, are low abundant and remain difficult to detect. Hence, these proteins are understudied and require larger sample sizes or meta-analyses^26^. Our *cis*-pQTL hits include proteins from a variety of biological processes and tissues such as the lymphoid cell surface receptor CD200R1, the blood secreted interleukins IL12B or IL18, the dendritic MHC complex class II DR alpha (HLA-DRA), placenta growth factor PDF, or the macrophage protein TREM2. Of the 135 novel *trans*-pQTLs identified, we successfully validated 54% using the eQTLGen whole-blood meta-analysis. Tier 1 (Validated Blood Regulators) showed a strong association with the expression of the nearest regulatory gene (e.g. CCDC77, ABHD5, AGK) suggesting a classic transcription-mediated mechanism. Notably, Tier 2 (Direct Target eQTL) hits (e.g. MGMT, TNFRSF11A, LGALS9) revealed a direct eQTL effect on the distal target gene, implying long-range chromatin interactions. DBS-based pQTL studies are still rare, possibly contributing to the novel *trans*-pQTLs described here.

Our study highlighted temporal elevation of LAP3 levels in DBS from recently infected subjects. As an aminopeptidase involved in immune regulation, LAP3 has previously been reported to increase following SARS-CoV-2 infection and/or vaccination^16,27^ as well as in other viral infections, such as generalized lymphadenitis and adenovirus infection^28^. These findings suggest that LAP3 is not exclusively specific to SARS-CoV-2 infection but has a broader role in the human response to various viral or bacterial pathogens, as shown in a recent pan-disease study^4^. The transient cross-sectional decrease post infection is a unique insight from this study, as it occurred in all donor samples collected within 3-6 weeks post confirmed infection. Compared to other immune markers with a more rapid decline, such as CCL2 or CXCL9^29^, the gradual decrease of LAP3 DBS levels made it possible for us to capture these in a study with an exposure-unrelated (random) sampling event. It is therefore reasonable to assume other immune markers with previous links to SARS-CoV-2 infections were missed due to steeper post-infection decline. To complement the knowledge on latencies reported for IgG or IgM, studies with tighter sampling events would be required.

Our study included mainly inflammation-related proteins, hence it may have missed other important pathways or pathologies, such as coagulation or metabolic phenotypes. Including additional proteins in the targeted or discovery-driven analysis in future studies could identify other, likely more persistent, proteomic changes following infection. In clinical reports enriched for severe phenotypes, consistent elevation of immune proteins over a longer time has been linked with adverse outcome^30^. In cross-sectional population surveys, however, the heterogeneity of disease states, impact of age-accumulated co-morbidities, and spread of sampling timepoints influence our ability to draw simpler conclusions. Other studies have shown that many inflammation-related proteins return to baseline levels within two weeks post infection^29^. Here non-hospitalized young adults (age < 30) from the BAMSE cohort were included, who were neither instructed to change their behavior during the study nor told to change their self-sampling plans due to an infection or current symptoms.

The characteristics of our study population also influenced the outcome. While the inclusion of informed participants representing a generally healthy, younger segment of the population is an interesting aspect, it also poses challenges for detecting large variance in proteins levels in a rather homogeneous group. The cohort largely followed national recommendations at the time, and most individuals received vaccination as soon as it became available for their age group in Sweden. This resulted in overlapping seroconversion periods, and limiting the formation of sufficiently large, clearly separated comparison groups. The coinciding vaccination and start of the third sampling phase, and the appearance of the fast-spreading Omicron variant posted challenges when disentangling the longitudinal differences between the three collection phases. Therefore, it remains challenging to fully distinguish these interconnected events and assign specific biological effects to each event.

Longitudinal population-based DBS home-sampling captured robust insights into the dynamic molecular events occurring during three pandemic years. Together with cohort data on genetic variation, clinical diagnostics and health history, monitoring health on a molecular level in the self-sampled DBS highlighted the importance of time-resolved phenotyping when stratifying individuals at risk, as pre-existing and persistent but also short-term alterations could influence long term health outcomes.

## Author contribution

AB performed experiments, analyzed data, and wrote the manuscript. SB collected data, analyzed data, and wrote the manuscript. MK analyzed data and wrote the manuscript. SKM analyzed data. AK analyzed data and wrote the manuscript. ZY analyzed data, AV processed samples, LD developed software and analyzed data. KC provided resources. AB, IK, ASM, SE and AL collected data and provided funding. BM provided resources and developed software. NR conceptualized study, provided funding, provided resources. EM conceptualized study, supervised the work, provided funding, and provided resources. JMS conceptualized study, supervised the work, provided funding, provided resources, and wrote the manuscript. All authors read and approved the final version.

## Acknowledgements

First, we thank all BAMSE participants and staff that have contributed to data collection throughout the years. We thank Olga Mukalova and Olof Beck for technical support, Matilda Dale, the Schwenk and Nilsson Labs members for analytical support and fruitful discussions. We acknowledge the support from the SciLifeLab infrastructure units for Affinity Proteomics in Stockholm and Uppsala, and the Autoimmunity and Serology Profiling unit. We thank the KTH node of Protein Production Sweden (PPS), a national research infrastructure funded by the Swedish Research Council), the Human Secretome Project at the Wallenberg Center for Protein Research, and everyone at the Human Protein Atlas (HPA). The authors acknowledge support from the National Genomics Infrastructure in Stockholm funded by Science for Life Laboratory, the Knut and Alice Wallenberg Foundation and NAISS/Uppsala Multidisciplinary Center for Advanced Computational Science for assistance with massively parallel sequencing and access to the UPPMAX computational infrastructure.Funding

The BAMSE study received supported from grants of the Swedish Research Council (2016-03086; 2018-02524; 2019-01060; 2020-02170; 2022-06340; 2024-03164); Forskningsrådet för hälsa, Arbetsliv och välfärd (2017-00526); Formas (2016-01646); Hjärt-Lungfonden; Region Stockholm (ALF project); and the Asthma and Allergy Research Foundation.

JMS received funding from the SciLifeLab National COVID-19 Research Program, which was financed by the Knut and Alice Wallenberg Foundation (2020.0182, 2020.0241) and SciLifeLab’s Pandemic Laboratory Preparedness program (VC-2022-0028). NR received funds from Sweden’s innovation agency Vinnova (2020-04451). BM received funds form The Erling Persson Foundation (20210125). We also acknowledge the tremendous support from the Knut and Alice Wallenberg Foundation for funding the Human Protein Atlas. This work was partially supported by the Wallenberg AI, Autonomous Systems and Software Program (WASP) funded by the Knut and Alice Wallenberg Foundation.

## Data availability

The datasets of the BAMSE study are not publicly available due to legal and ethical regulations. Upon reasonable requests, data supporting the findings of this study are available from the PI of the BAMSE cohort (Professor Erik Melén).

## Materials and Methods

### Study population and sampling

The study population included 808 participants from the Swedish Barn [Children], Allergy, Milieu, Stockholm Epidemiology (BAMSE) birth cohort. It is originally a population-based prospective cohort consisting of 4,089 children born between 1994 and 1996 in Stockholm, Sweden. The participants have been followed up with repeated clinical examinations and questionnaires. During the pandemic, the participants included in the presented study were invited to participate in a COVID-19 follow-up divided into four phases^31^. The current study includes data from the three first phases (Phase 1 from August to November 2020, Phase 2 from October 2020 to June 2021, and Phase 3 from October 2021 to May 2022) and consists of web-based questionnaires and self-collection of quantitative dried blood spot (DBS, Capitainer AB) samples from finger pricking at all three phases. Further, a clinical examination was conducted at the Stockholm South General Hospital during Phase 2^32,33^. The DBS samples were self-sampled at home in Phase 1 and 3 or self-sampled at the clinical examination in Phase 2 assisted by a nurse. The home-collected samples were mailed back using regular mail, and all DBS samples were stored at room temperature until analysis. Informed written consent was obtained from all subjects, and ethical permits were granted by the Regional Ethical Review Board in Stockholm (DNR 2016/1380-31/2) and Swedish Ethical Review Authority (DNRs 2020-02922, 2024-06052-02)^31^.

### Questionnaire-, clinical and registry variables

Data on current smoking was self-reported in Phase 1 to 3 questionnaires. The subjects self-reported symptoms of suspected or confirmed COVID-19 in each phase, with the Phase 1 questionnaire covering the start of the pandemic until the date of questionnaire 1, with Phase 2 covering August 2020 until the date of questionnaire 2, and Phase 3 questionnaire covering the entire pandemic until date of questionnaire 3. Among subjects which reported symptoms, follow-up questions regarding symptom duration were used to define long-term symptoms extending over 2 months and 3 months. From the questionnaires, we also extracted self-reported data on positive COVID-19 PCR- or antibody-tests. At the clinical examination of Phase 2, body mass index (BMI) and body fat percentage were measured using a bioelectrical impedance machine (Tanita MC 780 body composition monitor). Additional SARS-CoV-2 IgG serology in plasma collected during Phase 2 clinical examination was obtained from the Karolinska University Hospital Laboratory standard SARS-CoV-2 IgG serology test. Information about confirmed infection was obtained from SmiNet, which is a national register for notifiable transmittable diseases in Sweden, covering all registered positive PCR tests (Public Health Agency of Sweden, SmiNet, In Swedish. Available at: https://www.folkhalsomyndigheten.se/sminet). A SmiNet-case was defined as a positive entry date in SmiNet preceding the date of DBS filter paper sampling within each respective phase. Information on vaccination against COVID-19 was obtained by linkage to the obligatory National Vaccination Register. COVID-19 vaccination was defined as receiving a vaccination dose preceding the date of DBS filter paper sampling in each respective phase.

### DBS sample preparation

After visual inspection of each sample card to determine if at least one of the paper discs on the sampling device was correctly filled with blood, we randomly transferred one disc per subject and timepoint into flat bottom 96-well plates (VWR, art no. 734-2327) using Semi-Automated Puncher (710-1100, PH96, Pre-analytical Handler 96 for Capitainer®, Intema). All samples (Phase 1, 2 and 3) from the same participant were transferred to the same plate, and each plate was filled with 84 discs. We then added 100 µl buffer (1x Phosphate-buffered saline (PBS, Medicago) with 0.05% Tween 20 and protease inhibitor cocktail (#04693116001, Roche) to each well and the discs were incubated under gentle rotational shaking (170 rpm) for 60 minutes at room temperature. Following centrifugation, all volume was transferred into a PCR plate (#732-4828, VWR) and were then centrifuged for 3 min at 2100 x g (Allegra X-12R, Beckman Coulter Inc.). From this plate, we collected 70 µl supernatant into a new PCR plate, and the eluates were stored at −80°C prior to the analysis.

### Detection of anti-SARS-CoV-2 antibodies in DBS

SARS-CoV-2 related recombinant proteins and antibodies used as controls, either obtained from commercial providers or produced as described previously^34,35^, were covalently coupled to color-coded magnetic microspheres (MagPlex, Luminex Corp.) using NHS/EDC chemistry as described elsewhere^13,16,36^. See antigen and antibody list in Tab.S2. To summarize, 4 µg proteins or 1.7 µg antibody were diluted in MES buffer [100 nM 2-(N-morpholino) ethanesulfonic acid (pH 5.0)] to a final volume of 100 µl. The carboxylated surface of the beads were then activated by incubating beads with 50 µl activation solution (N-hydroxysuccinimide (Pierce) and 1-ethyl-3-(3-dimethylaminopropyl)-carbodiimide (ProteoChem)) for 20 min. After washing with MES buffer, we added the diluted proteins to the beads and let them incubate for 2 h at 23°C. The beads were then washed with PBS-T 0.05% and stored in Blocking Buffer for ELISA (BRE, 11112589001, Roche) overnight, before we pooled beads of different bead identities to form a suspension bead array (SBA).

On the day of the assay, the blood eluates were thawed at 4°C and diluted 1:2.5 in assay buffer containing 1x PBS with 0.05% Tween20, 3% BSA (B2000-500, Lot# 04E6171, Saveen Werner) and 5% milk powder (70166-500 G, Lot# 102586039, Sigma-Aldrich). We then transferred 35 µl of the diluted eluates to 384-well plates containing 5 µl antigen SBA, and incubated the plates in the dark for 60 min at 23°C, shaking at 650 rpm. Subsequently, the plates were washed three times with 60 µl PBS-T 0.05% using an automated washing system (Biotek EL406), and 50 µl anti-human IgG-R-PE (H10104, Lot#2306068, Invitrogen) diluted to 0.4 µg/ml in PBS-T 0.05% was added to the beads. The beads were then incubated for 30 min at 23°C, shaking at 650 rpm. in the dark. After three final washes with PBS-T 0.05%, we added 60 µl PBS-T 0.05% to the beads, and the fluorescence associated with each bead was measured in a FlexMap 3D instrument (Luminex Corp.). Signals were reported as median intensity fluorescence (MFI) per antigen and sample, where each data point had a bead count of at least 32 per ID.

Serological classification of subjects was determined by a threshold set at 6x SD over the peak of the population density of the first two sampling phases.

### Autoreactive antibody assays in DBS

In addition to the COVID-19 antigens, 21 human interferons (IFNs) were included in the bead mixture of the SBA. The IFNs were produced as described elsewhere^37^. The protein coupling, analysis and readout was performed as described for the detection of SARS-CoV-2 antibodies described above.

### Affinity proteomics assays in DBS

To measure relative protein levels in the samples, the samples were analyzed using the Olink Explore 384 Inflammation panel (Lot # B23405, Olink) at the SciLifeLab Affinity Proteomics Unit in Uppsala, Sweden. In short, 1 µl eluate is incubated with antibodies labelled with unique DNA oligonucleotides. If the two matched antibodies simultaneously bind to a target protein, their DNA tags can hybridize, and polymerase-dependent extension is used to create a double-stranded DNA ‘barcode’ unique for the specific antigen, and to amplify the signal. The resulting amplicon is then quantified by Next Generation Sequencing readout on Illumina instruments, and the reported Ct values are converted to Normalized Protein eXpression (NPX) values using Olink NPX Explore Software (version 3.0.5) and Intensity normalization. The NPX values are presented as arbitrary units (AU) in the log2 scale. In total, 365 inflammation-associated proteins were measured in 2398 of the samples.

### Complete blood cell counts

Complete blood counts (CBC) were assessed at Phase 2 of the BAMSE COVID-19 follow-up and at the BAMSE 24-year follow-up using whole blood collected via venipuncture. The 24-year follow-up took place between November 2016 and May 2019, approximately 1-4 years before Phase 1 of the COVID-19 follow-up (starting in August 2020). Blood count measurements were conducted by routine flow cytometry at Karolinska University Laboratory in Stockholm, Sweden, with reference values provided by the same laboratory^18,19^. Fig. S11 presents the distributions of CBC parameters at Phase 2, based on data from 782 subjects with complete CBC and Olink protein data from Phase 2 of the BAMSE COVID-19 follow-up. Given the pronounced skewness of basophil count toward zero, this parameter was excluded from downstream analyses.

### Affinity proteomics assays in plasma

At the BAMSE 24-year follow-up, the relative levels of 92 inflammation-related plasma proteins were assessed using the Olink Target 96 Inflammation panel (Olink Biosciences, Uppsala, Sweden). Plasma was obtained from venous blood collected in EDTA tubes. The analytical method was similar to the method used for Olink Explore 384 Inflammation panel, except for using quantitative real-time polymerase chain reaction instead of next-generation sequencing. Samples that deviated more than 0.3 NPX from the median value of an internal control were excluded.

### Statistical analyses

All data analyses and visualizations were performed using R (version 4.4.1) programming language^38^ or STATA SE 19.5 unless otherwise stated. Tidyverse (version R 2.0.0)^39^ was used for data processing and analysis, and plots were created using the ggplot2 package (version 3.5.1)^40^, together with the patchwork (version 1.3.0)^41^, RColorBrewer (version 1.1-3)^42^, ggh4x (version 0.3.1)^43^, ggbeeswarm (version 0.7.2)^44^, ggrepel (version 0.9.5)^45^, ggsignif (version 0.6.4)^46^, GGally (version 2.4.0)^47^, ComplexHeatmap (version 2.18.0)^48^, viridis (version 0.6.5)^49^, Ghibli (version 0.3.4)^50^, ggalluvial (version 0.12.5)^51^. The psych-package (version 2.4.6.26)^52^ was used for calculating Cohen’s kappa and intra-class correlations (ICC).

### Demographic data

For demographic data displayed in Tab. 1 and 2. Fisher’s exact test was used to analyze dichotomous variables which are displayed as number of subjects and percentage, and the Mann-Whitney U-test or Kruskal-Wallis test was used to analyze continuous variables which are displayed as medians with range.

### Serological data analysis

Serological assay data were normalized using a mixed model implemented in Julia (version 1.7.2)^53^ programming language to remove correlation with a negative control bead for all analytes, except for spike-and RBD-based versions, which were normalized against an anti-human IgM bead to account for rising antibody levels post-vaccination. Data from all sampling phases were normalized together for all analytes except for the interferons, which were normalized per sampling phase. The model, applied to log2-transformed data as previously described^13,16^, incorporated both linear and uniform distribution components to account for distributions where most samples exhibit low baseline levels, while a subset displayed higher positive values. The resulting relative log2 values were scaled and centered per protein to put them on the same scale.

Seropositivity for each anti-SARS-CoV-2 antibody was determined using a population-based threshold of 6 standard deviations (SD) of the estimated negative proportion above the gaussian population peak as previously described^13^. The negative population consisted of unvaccinated individuals without PCR-confirmed infection. This threshold, established using data from sampling Phases 1 and 2, was applied to Phase 3. Phase 3 was excluded from threshold determination due to the high proportion of vaccinated individuals in this phase. To determine the number of subjects who remained seronegative across all three sampling phases, serostatus was first determined at the sample level. For each sample, the antigen-specific seropositivity was summarized separately for N, S, and RBD by applying a majority rule across all corresponding antigens within each group. A sample was then classified as seropositive if at least two of the three antigen groups (N, S, RBD) were seropositive. Subjects were considered seronegative across all phases if all samples collected from that subject were classified as seronegative.

Cohen’s kappa coefficient was calculated using the psych R package (version 2.4.6.26)^54^, to quantify the level of agreement between the seropositivity estimated in this study and seropositivity estimated using ELISA assays targeting the Spike protein on plasma samples from sampling Phase 2. For seropositivity determination against interferons, signals were transformed using robust z-scores per phase, and a cutoff of 3.5 was applied. Hierarchical clustering, using maximum distance and ward.D2 agglomeration method, was performed on six binders (N full length, NC, S1, S-WT, RBD-WT, and RBD Omicron) to stratify samples into four serocluster groups based on serological profiles. These seroclusters and the seropositivity for each antibody were then compared to available vaccination and infection status data.

### Proteomics data analysis

Intensity normalized protein measurements and Plate control normalized protein measurements were obtained from Olink. In general, we used the intensity normalized data for downstream analysis since this normalization method gives more stable NPX values for proteins not highly expressed in the Olink-specific plate controls.

To minimize sample-to-sample variation due to sample handling and non-specific protein correlation, the data were normalized using the ProtPQN method with the ProtPQN R package (version 1.0.1)^55^. Only samples from the BAMSE cohort were included in the ProtPQN normalization.

Linear models were performed using the stats base R package (4.4.1)^38^. For protein associations between age, sex, BMI, and smoking, we fitted linear models for all samples from Phase 2 for each of the proteins against each of the variables. P-values were adjusted using FDR-correction, and were, together with the slopes, compared to the slopes and Bonferroni-adjusted p-values from the UK biobank (UKB) data^1^ and previous DBS data^16^ for the overlapping proteins and variables. For the BAMSE data, we used p-value < 0.05 as significance levels, whereas the UKB data set used p < 1.7e-5.

To assess protein stability over time, we calculated Intra-class Correlation Coefficients (ICCs) for each protein using the ICC() function from the R psych^54^ package. Out of the various ICC forms available, we specifically chose the ICC(3,1) model. We opted for ICC(3,1) because it represents a two-way mixed-effects model for absolute agreement of single measures. Proteins were deemed stable if their calculated ICC(3,1) value was ≥0.75, based on suggested guidelines on the interpretation of ICC values^56^.

Linear mixed-effect models (LMMs) were performed using the *lmerTest* R package (version 3.1-3)^57^. LMMs were applied to evaluate associations between protein levels and storage time among uninfected and unvaccinated individuals across all three sampling phases, adjusting for sex and including a random intercept for each subject. This analysis was performed on 25 subjects who consistently remained in Serocluster 1 across all three sampling phases, minimizing the potential influence of infection or vaccination on protein levels. We then assessed the enrichment of storage-sensitive proteins across protein classes (predicted secreted, intracellular, and/or membrane proteins) from Human Protein Atlas using Fisher’s exact test.

To compare protein levels between subgroups of the data to investigate the effect of infection and vaccination, the data was first adjusted for storage time by taking the residuals from a LMM between protein levels and storage time with random slope per subject. Using the data adjusted for storage time, we first compared naïve seronegatives (Serocluster 1) who got infected prior to Phase 2 (Serocluster 2; n = 96) to a randomly sampled group of naïve who remained naïve (n = 96) bootstrapped over 100 iterations, and to infected who remained infected (Serocluster 2, n = 77). We also compared the randomly sampled groups of naïve who remained naïve to the subjects who remained infected.

Differences between protein levels between groups within each sampling phase, as well as the median changes over time between the groups, were assessed using linear models adjusted for sex using the lm function from the *stats* R package. P-values were adjusted using FDR correction method using the p.adjust function in *stats*. When comparing two groups of unequal sample sizes, the same number of subjects as in the smaller group were bootstrapped from the larger group with replacement for 100 iterations, and the statistical comparison was performed in each iteration. The results from the bootstrapped analyses were summarized using the median effect size and corresponding estimates across all iterations, with FDR control applied to the aggregated results.

Paired Wilcoxon test was used to compare protein levels before and after PCR-confirmed infection (N=55), using R function wilcox.test from the stats package. First, only infected subjects who were not vaccinated were included in the analysis, and the post-infection groups were divided into two time-interval groups (0-30 days (N=12) and 30+ days (N = 43)) to capture non-linear temporal changes. Non-paired Wilcoxon test was used between the two time-interval groups since they did not share any subjects. In a second comparison, samples from subjects who had gotten vaccinated before PCR-confirmed infection were also included in the analysis, and an unpaired Wilcoxon test was used to compare pre-infection samples (N=203 samples, 133 unique subjects), and samples from the two time interval groups (0-30 days (N=23) and 30+ days (134 unique subjects, and 201 samples) against each other. As a control, we included subjects who only got vaccinated and not infected, so we excluded samples from Serocluster 2 and 3 which contained infected subjects with antibodies against nucleocapsid proteins. We took the most recent sample following a vaccination, and the most recent sample pre-vaccination for each subject and compared these with a paired Wilcoxon test. Comparison between the two time-intervals post vaccination was made using unpaired Wilcoxon test since these samples were not from the same subjects.

For plotting protein levels in relation to time since infection, all proteins levels were first transformed into z-scores. Then all samples from subjects with a positive PCR test were extracted. Samples collected after vaccination were then excluded, and the remaining samples were matched to have one pre and one post infection sample per subject. In total, 110 samples from 55 subjects were included. The date for PCR-confirmed infection was used as the infection date, acknowledging that this does not perfectly correspond to the true date of infection due to the delay between infection and testing. For plotting protein levels in relation to time since vaccination, matched pre-and post-vaccination samples closest in time to the vaccination date were selected. Samples from subjects belonging to infected seroclusters 2 or 3 were excluded. In total, 950 samples from 475 subjects were included.

Fisher’s exact test was used to compare the number of samples above z-score = 2 or below z-scores = −2 for the infected and vaccinated groups described above, using R function fisher.test function from the *stats* R package. Extracted p-values were FDR corrected.

### Genome-wide association of proteomics data

To further understand the genetic architecture of these protein levels in DBS, we performed a protein quantitative trait loci (pQTL) analysis. The Phase 1 Olink data was used for this analysis due to its limited influence by confounders such as infections and vaccination. The genotyping and imputation protocols for the BAMSE samples have been previously described ^58^. Basically, genotyping for the full cohort (n≅2892) was performed in two distinct waves, separated by approximately ten years, utilizing different Illumina platforms (Illumina Human 610-quad BeadChip and Illumina Infinium Global Screening Array-24 v1.0 (GSA) chips). Standard quality control measures were applied independently to the assays in both waves for genotype and sample quality control, imputation quality control (QC) that were anchored to the HRC reference panel.

Of the 808 samples with Olink proteomic data, we successfully matched 678 samples to the imputed data. Following QC for available covariates, which resulted in the exclusion of n=28 samples, the final analysis set comprised of 650 samples. The ProtPQN intensity normalized NPX values for each Olink protein from sampling Phase 1 was inverse rank normalized and adjusted for gender, age, and top 20 genetic principal components (PCs). To leverage the total cohort size, a Genome-Wide Association Study (GWAS) was first performed independently for each of the 365 Olink-measured inflammation-related DBS proteins in Wave1 (n=95 samples) and Wave2 (n=555 samples), using Plink 2.0 on chromosomes 1-22, on all biallelic SNPs filtered at MAF > 0.01 and imputation quality > 0.3 in each wave. The summary statistics from both the waves were subsequently combined via a fixed-effects inverse variance-weighted (IVW method) meta-analysis implemented in the METAL software^59^, to generate the final pQTL estimates for the combined waves. This approach ensured that the genetic effects were consistently estimated across the two genotyping waves. The meta-analysis was restricted to variants that were present in both the waves. The statistical significance was defined as P ≤ 1 × 10^−10^ (using Bonferroni corrected GWAS threshold of P ≤ 5 × 10^−8^ for 365 proteins). Using GCTA-COJO method, conditionally independent association signals within a locus were identified by feeding the LD information generated from Plink1.9 on wave2 imputed SNPs. When the SNP’s position was within a 1Mb window of the protein’s gene, these pairs were classified as *cis*-pQTLs, and those outside that window were classified as *trans*-pQTLs. We then checked for overlap with prior reported pQTLs among primarily European populations in UKB^1^, SCALLOP^26^, OpenGWAS^60^, and GPPAD^21^. The eQTLGen SIGNIFICANT RESULTS (2019-12-11 Release) was used to annotate the novel *trans*-pQTLs.

### Analyses of blood cell counts and circulating DBS proteins

We first applied Spearman correlation to study associations between DBS Olink proteins and CBC parameters in 782 subjects with complete protein and CBC data from Phase 2 of the COVID-19 follow-up. Hierarchical clustering was performed on both CBC parameters and DBS proteins using the “average” method on a distance matrix derived from the Spearman correlation coefficients.

Secondly, we examined correlations between 51 proteins measured in both DBS (COVID-19 follow-up) and plasma (24-year follow-up). The analysis included 690 subjects from the 24-year follow-up with complete CBC and protein data at both time points; 782 subjects were included from Phase 2. Lastly, we examined the relationship between CBC parameters and plasma proteins at the 24-year follow-up employing Spearman correlation, and compared these results to the corresponding correlations for the same 51 proteins measured in DBS. Shared protein-level associations across CBC parameters were evaluated via hierarchical clustering (average linkage) using correlation-based distance metrics. Statistical significance was defined as p < 0.05 after false discovery rate (FDR) correctio. All analyses were performed in RStudio version 4.4.1 (2024-06-14)^38^, and visualizations were created using the R packages ggplot2^40^, grid^38^, gridExtra^61^, pheatmap^61^, rcartocolor^62^, and UpsetR^63^.

